# Not all mosquitoes are created equal: incriminating mosquitoes as vectors of arboviruses

**DOI:** 10.1101/2022.03.08.22272101

**Authors:** Morgan P. Kain, Eloise B. Skinner, Tejas S. Athni, Ana L. Ramirez, Erin A. Mordecai, Andrew F. van den Hurk

**Affiliations:** Department of Biology, Stanford University, Stanford, CA, 94305, USA; Natural Capital Project, Woods Institute for the Environment, Stanford University, Stanford, CA 94305, USA; Centre for Planetary Health and Food Security, Griffith University, Gold Coast, QLD, 4222, Australia; Department of Pathology, Microbiology, and Immunology, University of California, Davis, Davis, CA 95616; Public Health Virology, Forensic and Scientific Services, Department of Health, Queensland Government, Brisbane, Queensland, Australia

**Keywords:** Mosquito, arbovirus, vector competence, public health, Australia, infection barriers

## Abstract

The globalization of mosquito-borne arboviral diseases has placed more than half of the human population at risk. Understanding arbovirus ecology, including the role individual mosquito species play in virus transmission cycles, is critical for limiting disease. Canonical virus-vector groupings, such as *Aedes*- or *Culex*-associated flaviviruses, have historically been defined using phylogenetic associations, virus isolation in the field, and mosquito feeding patterns. These associations less frequently rely on vector competence, which quantifies the intrinsic ability of a mosquito to become infected with and transmit a virus during a subsequent blood feed. Herein, we quantitatively synthesize data from 80 laboratory vector competence studies of 115 mosquito-virus pairings of Australian mosquito species and viruses of public health concern to further substantiate existing canonical vector-virus groupings, uncover new associations, and quantify variation within these groupings. Our synthesis reinforces current canonical vector-virus groupings but reveals substantial variation within them. While *Aedes* species were generally the most competent vectors of canonical “*Aedes*-associated flaviviruses” (such as dengue, Zika, and yellow fever viruses), there are some notable exceptions; for example, *Aedes notoscriptus* is an incompetent vector of dengue viruses. *Culex* spp. were the most competent vectors of many traditionally *Culex*-associated flaviviruses including West Nile, Japanese encephalitis and Murray Valley encephalitis viruses, although some *Aedes* spp. are also moderately competent vectors of these viruses. Conversely, many mosquito genera were associated with the transmission of the arthritogenic alphaviruses, Ross River, Barmah Forest, and chikungunya viruses. We also confirm that vector competence is impacted by multiple barriers to infection and transmission within the mesenteron and salivary glands of the mosquito. Although these barriers represent important bottlenecks, species that were susceptible to infection with a virus were often likely to transmit it. Importantly, this synthesis provides essential information on what species need to be targeted in mosquito control programs.

**Author summary:** There are over 3,500 species of mosquitoes in the world, but only a small proportion are considered important vectors of arboviruses. Vector competence, the physiological ability of a mosquito to become infected with and transmit arboviruses, is used in combination with virus detection in field populations and analysis of vertebrate host feeding patterns to incriminate mosquito species in virus transmission cycles. Here, we quantified the vector competence of Australian mosquitoes for endemic and exotic viruses of public health concern by analyzing 80 laboratory studies of 115 mosquito-virus pairings. We found that Australia has species that could serve as efficient vectors for each virus tested and it is these species that should be targeted in control programs. We also corroborate previously identified virus-mosquito associations at the mosquito genus level but show that there is considerable variation in vector competence between species within a genus. We also confirmed that vector competence is influenced by infection barriers within the mosquito and the experimental protocols employed. The framework we developed could be used to synthesize vector competence experiments in other regions or expanded to a world-wide overview.

## Introduction

Mosquito-borne arboviruses infect hundreds of millions of people and animals annually thus representing a major global public and veterinary health threat [1, 2]. The viruses responsible for severe clinical disease predominately belong to the viral families *Flaviviridae* and *Togaviridae* [2]. Globally important flaviviruses include the dengue viruses (DENVs) which cause 50 to 100 million symptomatic infections each year, the encephalitogenic Japanese encephalitis (JEV) and West Nile (WNV) viruses, which have expanded their geographical ranges in the last 25 years, and Zika virus (ZIKV), which is now recognized as a potential cause of severe congenital abnormalities [3-5]. Alphaviruses in the family *Togaviridae* from the Americas, such as eastern, western and Venezuelan encephalitis viruses are associated with encephalitis, whilst chikungunya (CHIKV) and Ross River (RRV) viruses, which originated in Africa and Australasia, respectively, have caused explosive epidemics of debilitating polyarthritis as they have invaded virgin ecosystems [6].

Combating mosquito-borne disease begins with understanding which mosquito species can transmit these viruses and how effectively they do so. Broad and extensive knowledge regarding the importance of different mosquito genera as arbovirus vectors has been gained in previous studies through phylogenetic analyses of the genetic groupings of arboviral families and their correlation with known associated mosquito genera. Phylogenetic analysis of non-structural five (NS5) gene sequences coupled with their respective vector associations initially classified the flaviviruses into three clusters: viruses without a known vector, tick-borne viruses, and mosquito-borne viruses [7]. Further analyses by Gaunt et al. [8] and Moureau et al. [9] using additional genes and more comprehensive datasets subdivided the mosquito-borne flaviviruses into *Aedes*-associated flaviviruses (AAFVs; e.g., DENVs in *Ae. aegypti* [10]) and *Culex*-associated flaviviruses (CAFVs; e.g., WNV in members of the *Culex pipiens* species complex [11]). Isolation of viruses from mosquitoes in the field and identification of mosquito host-feeding patterns, specifically whether vector species feed on vertebrate hosts of the virus [8], has been used to reinforce these classifications.

Groupings based on these criteria can only provide an indirect or inferred measure of the importance of different mosquito genera as arbovirus vectors. Assessing the innate physiological ability of each mosquito species to act as a vector can strengthen mosquito genus–viral family groupings and help to quantify variation within them. Laboratory-based vector competence studies provide quantitative measures of a mosquito’s physiological ability to become infected with and transmit arboviruses by assessing the progression of a pathogen through four different barriers primarily associated with the mosquito mesenteron (midgut) and salivary glands (Fig 1), all of which must be overcome for the virus to ultimately be transmitted to a susceptible host [12-15]. After a mosquito takes a virus-laden blood meal, the virus must first overcome the mosquito’s mesenteronal infection barrier by binding to specific receptors on the epithelial cells (Fig 1, part 2). Passage of this barrier is measured by detecting virus in an infected mosquito’s body or the dissected mesenteron. The pathogen must then bypass the mesenteronal escape barrier, which limits dissemination of the pathogen from the mesenteronal epithelial cells [16, 17] (Fig 1, part 3). The detection of virus in the legs, wings or head of a mosquito indicates that the virus has disseminated from the mesenteron via the hemolymph to other tissues and organs (Fig 1, part 4; [18]). Finally, the pathogen must successfully travel to, infect and replicate within, the salivary glands to a level sufficient for transmission [14, 19-21] (Fig 1, parts 5-6). Alternatively, if the virus does not infect the salivary glands or is not released in the saliva during probing and/or feeding, then the mosquito is considered to possess salivary gland infection or escape barriers, respectively. Expression of one or more of these barriers ultimately limits the ability of a given mosquito to transmit a virus.

**Figure 1.**
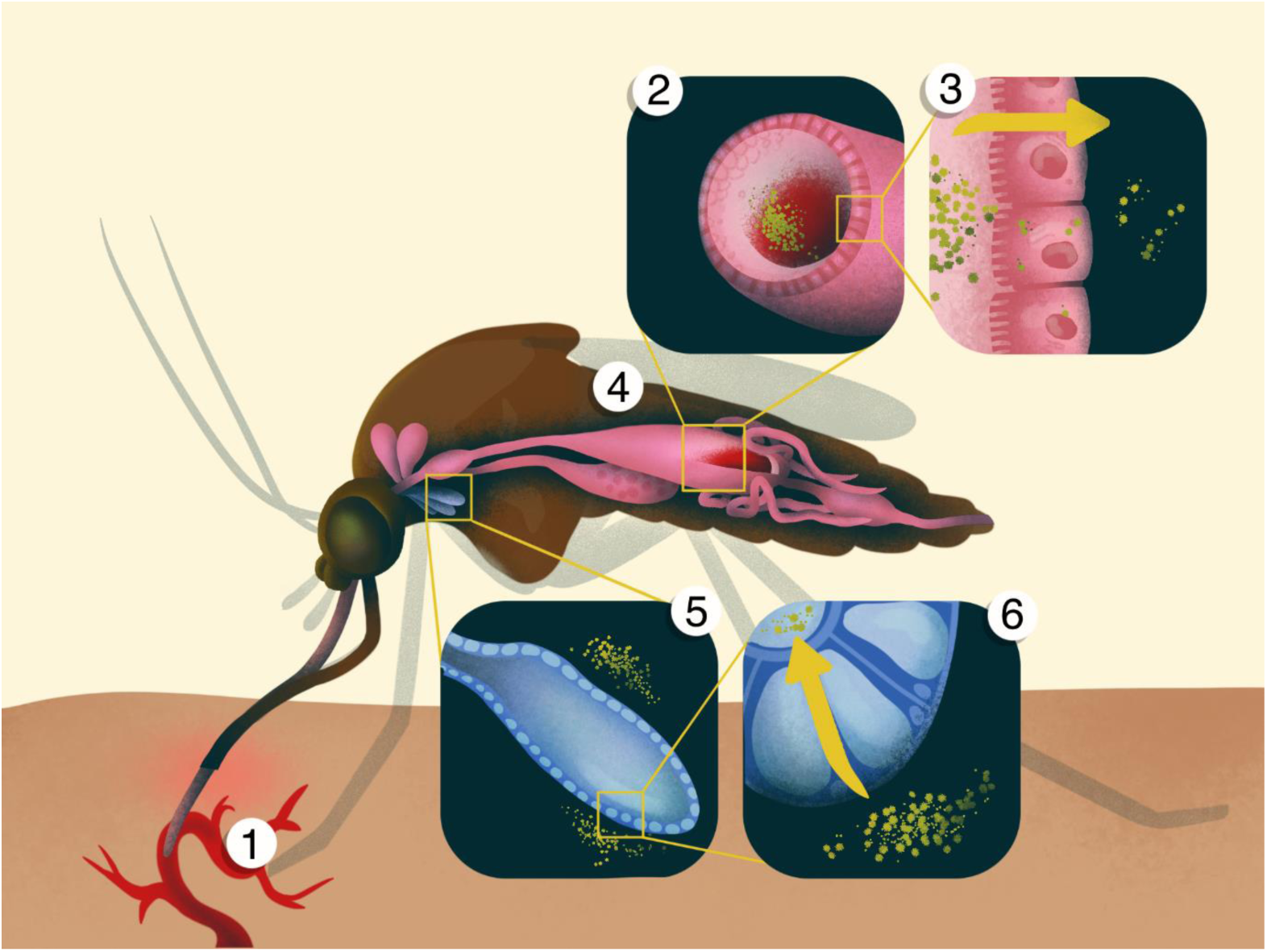
The steps involved in the successful transmission of an arbovirus by a mosquito. 1. Virus is ingested along with the blood meal from a viremic host; 2. The infected blood meal is deposited in the posterior section of the mesenteron (midgut). 3. The virus infects and replicates in the epithelial cells of the mesenteron; 4. The virus disseminates from the mesenteronal epithelial cells via the hemolymph and infects other tissues, such as fat bodies or neural tissue, where it can undergo another round of replication. 5. The virus then infects the cells of the salivary glands. 6. The virus is then released in the saliva when the mosquito probes another vertebrate host.

In vector competence experiments, mosquitoes are exposed to either a previously infected animal or an artificial blood meal containing the virus [22]. At different time points post-exposure, mosquito infection and disseminated infection status is tested by processing whole bodies or body parts, such as mesenterons, heads, legs and/or wings. Mosquitoes are then assessed for their ability to transmit the pathogen using either a susceptible animal model [18, 23] or an *in vitro* system of saliva collection [24]. Mosquitoes that do not express barriers to infection, dissemination, and transmission from the salivary glands are considered competent vectors. Conversely, species that strongly express any barriers that limit transmission are considered either poorly competent or incompetent; species whose mesenterons do not get infected after being exposed to an infectious blood meal are refractory to infection.

As single experiments tend to focus on the competence of specific mosquito species for a specific virus, a synthesis of many vector competence experiments is needed to corroborate known, identify new, and quantify variation within virus–mosquito associations across a broad range of mosquito and virus species. Australia is a particularly interesting case study for assessing vector competence because the island continent is climatically and biologically diverse, contains many ecologically distinct native and introduced mosquito species, and has experienced historical and emerging arbovirus threats. Australia has a history of outbreaks of endemic and exotic mosquito-borne arboviruses, the latter initiated by arrival of viremic travelers [25-28], and is potentially receptive to transmission of globally emergent arboviruses, such as ZIKV, CHIKV and the highly pathogenic North American WNV strains [29-31]. Thus, numerous laboratory-based experiments have been undertaken to assess the vector competence of Australian mosquito species for a range of virus species, with the overall aims of incriminating vectors and aiding targeted control strategies. Using data and measures of vector competence from 80 publications on Australian mosquito species, this study broadly assesses patterns of vector competence between mosquito genera and medically important viruses of the *Togaviridae* and *Flaviviridae* families. Importantly, by focusing on a single geographical region where common mosquito species have been tested for a range of endemic and exotic viruses, we avoid the issues of using data on a global scale, which is disproportionately centered on a limited number of mosquito species and viruses that present the largest public health threats. The Australian vector competence studies provide the ideal data for us to address four primary sets of questions:

1. Do mosquito–virus associations derived from Australian vector competence experiments reinforce or refute canonical associations based on phylogenetic relationships, virus isolation studies, and analysis of host feeding patterns? For example, are the canonical groupings of *Aedes*- or *Culex*-associated flaviviruses supported at the genus level, and are AAFVs and CAFVs only transmitted by *Aedes* spp. and *Culex* spp. respectively?
2. How much variation exists among mosquito species within a genus in their competence for the viruses within a given group?
3. What is the strength of the correlation between infection, dissemination, and transmission and which barriers to these processes are most limiting for vector competence?
4. What implications do the experimental design, methodology and extensiveness of individual studies have, and how does this affect our understanding of vector competence?

## Methods

### Data collection

All available Australian vector competence studies were collected by a single author (AvdH) using electronic databases, reference lists, hand searches, and institutional reports. A second author (EBS) then systematically searched for additional publications using a combination of the following search terms on Google Scholar: (mosquito* OR vector*) AND (competence OR infection OR dissemination OR transmission) AND (virus OR arbovirus OR flavivirus OR alphavirus) AND Australia. Studies were included if: a) they were original research; b) the viruses used were of human health importance; c) they assessed at least one mosquito species with an Australian genetic background. These studies were then filtered to ensure that: a) viruses originated from field isolates and were not derived from infectious clones or recombinants (i.e. [32]); b) mosquitoes had not been modified so that virus replication was impacted (i.e. *Wolbachia* transinfected [33]); c) mosquitoes were exposed to virus by feeding on a viremic animal or an artificial infectious blood meal (i.e. not by intrathoracic inoculation [34], whereby the mesonteronal infection and escape barriers are bypassed); d) all mosquitoes were tested individually (i.e. transmission was not measured in batches); e) a minimum sample size of five mosquitoes at the experimental endpoint for each mosquito-vector pair; and f) transmission was measured via feeding on a suitable animal model or detection in saliva expectorates collected using *in vitro* saliva collection methods (as opposed to detection of virus in salivary glands which does not take into account the presence of a salivary gland escape barrier to transmission). In total, we gathered data from 80 publications, which included 295 individual experimental treatments (i.e., the exposure of a single cohort of mosquitoes of a given species to a given virus in a set of experimental conditions) on more than 25,000 total individual mosquitoes across 27 species spanning 6 genera and 13 viruses spanning 2 families (counting the four dengue virus serotypes separately) (Table 1). All data are available in the S1 File. The following data were extracted from each study:

**Table 1.** Arboviruses assessed in vector competence experiments involving Australian mosquito species and included in the analysis.

| **Virus family** | **Virus species** | **Geographical distribution** | **Disease syndrome<sup>a</sup>** | **Vertebrate hosts<sup>b</sup>** | **Associated mosquito genera<sup>c</sup>** |
| --- | --- | --- | --- | --- | --- |
| *Flaviviridae* | Dengue types 1-4 | Asia, Africa, South and Central America | Haemorrhagic fever | Humans, non-human primates | *Aedes* |
|  | Yellow fever | Africa, South America | Haemorrhagic fever | Humans, non-human primates | *Aedes*,<br>*Haemagogus*,<br>*Sabethes* |
|  | Zika | Africa, Asia, South and Central America | Congenital malformations, Guillain-Barré syndrome | Humans, non-human primates | *Aedes* |
|  | Japanese encephalitis | Asia, Australasia | Encephalitis, meningitis | Birds, Pigs | *Culex* |
|  | Murray Valley encephalitis | Australasia | Encephalitis | Birds | *Culex* |
|  | West Nile<sup>d</sup> | Africa, Asia, Australasia,<br>North and South<br>America, Europe | Encephalitis,<br>meningitis | Birds | *Culex* |
| *Togaviridae* | Barmah Forest | Australasia | Arthritis | Mammals, birds | *Aedes, Culex,*<br>*Coquillettidia,*<br>*Verrallina* |
|  | Chikungunya | Africa, Asia, South and<br>Central America | Arthritis | Humans, non-human primates | *Aedes* |
|  | Ross River | Australasia | Arthritis | Marsupials, humans | *Aedes, Culex,*<br>*Coquillettidia,*<br>*Verrallina* |

<sup>a</sup>Many arbovirus infections cause non-specific febrile illness; the syndromes presented are the severe manifestations of disease.
<sup>b</sup>These are the major vertebrate hosts associated with transmission.
<sup>c</sup>Association of mosquito genera with the virus is based on virus isolation from the field and interaction with the vertebrate host as based on blood feeding patterns
<sup>d</sup>Includes the highly pathogenic North American strain and the Kunjin subtype which occurs in Australasia.

- Vector characteristics: Mosquito species; origin
- Arbovirus characteristics: Virus species; isolate or strain
- Study methods: Virus dose to which mosquitoes were exposed; unit of measure of viral titer of the infectious blood meal (such as plaque forming units (PFU), tissue culture infectious dose_50_ (TCID_50_), suckling mouse intracerebral inoculation lethal dose_50_ (SMIC ID_50_) which were all converted to infectious units per milliliter (IU/mL) for the analysis); method of virus exposure; method used to demonstrate transmission; assay to detect evidence of infection and/or transmission; days post exposure (DPE) to virus that infection, dissemination, and transmission were assessed; temperature at which mosquitoes were maintained; total number of mosquitoes exposed to the virus that were tested for infection, dissemination, and transmission
- Study results: proportion of mosquitoes that were exposed to the virus that were positive for infection, dissemination, and transmission

### Vector competence metrics and analysis

#### Maximal proportion infected, disseminated, and transmitting

Across each unique mosquito species–virus pairing assessed in the vector competence experiments, we extracted the highest observed proportion of mosquitoes of a given species that became infected with a given virus after ingestion of an infectious blood meal, irrespective of the dose administered or the day tested. We similarly identified the highest observed proportion of mosquitoes that developed a disseminated infection and that transmitted the virus. For each mosquito-virus pair, these maximal measures captured the single highest proportion observed across all viral doses the mosquito species was exposed to and all the days on which the species was tested. We chose this maximal measure, instead of a metric such as an average, to ensure that potentially meaningful mosquito-virus associations were not diluted by larger numbers of mosquitoes infected at low doses or tested at early time points post-exposure. Due to destructive sampling, experimentally observed proportions of mosquitoes positive for infection, dissemination, and transmission are conditioned on exposure (i.e., transmission is not conditioned on those already testing positive for dissemination).

For each virus, infection data were available for at least three species of mosquitoes (median of 7.5 species per virus) and dissemination data for at least two mosquito species per virus (median of 4 species per virus). Whilst transmission by individual mosquitoes was not assessed for some viruses (i.e. dengue virus type 1 (DENV-1)), the median number of species tested for transmission capability per virus was higher than for both infection and dissemination (median of 9 species per virus).

We statistically tested for differences in maximal infection proportion and maximal transmission proportion among the mosquito genera in our data set (*Aedes, Anopheles, Coquillettidia, Culex, Mansonia*, and *Verrallina*) for the following virus groups: *Aedes*-associated flaviviruses (AAFVs; yellow fever [YFV], ZIKV, DENV 1-4; *Culex*-associated flaviviruses (CAFVs; JEV, West Nile New York strain [WNV_NY99_], West Nile Kunjin strain [WNV_KUN_], Murray Valley encephalitis [MVEV]); and arthritogenic alphaviruses (CHIKV, RRV and Barmah Forest virus [BFV]). To do so we fit two regression models, each with a Binomial error distribution, that used the maximum detected proportion of mosquitoes infected or transmitting as the response variable; for weights (sample size) we used the number of mosquitoes included in the experimental treatment(s) in which the maximum proportion infected or transmitting was found. Each model included the following fixed effects: an interaction of mosquito genus and virus group, experimental dose, the number of DPE that mosquitoes were tested for infection or transmission, and the total mosquitoes exposed across all experimental infections as a measure of total research effort. Given our interest in identifying differences in the ability of mosquito genera to become infected with and transmit different viral groups, each unique mosquito species–virus pair within a broader mosquito genus–virus group pairing was considered a replicate for that mosquito genus–virus group. If more than one experimental treatment for a given mosquito-virus pair found an equivalent maximum, we summed the number of mosquitoes across the experimental treatments and used the average of virus titer the mosquitoes were exposed to and the number of DPE that the mosquitoes were tested to keep the total number of replicate data points for all mosquito genus–virus group pairings equal to the total number of unique combinations of mosquito species exposed to viruses within that virus group.

We fitted these regression models using the *glm* function in R [35] using the penalized maximum likelihood estimation procedure *brglmFit* from the *brglm2* package [36]. This procedure implements a quasi-Fisher scoring iteration [37] to allow for the estimation of confidence intervals (CI) in the presence of complete separation (groups with all zeros) [38] (e.g. all *Culex* mosquitoes tested negative for transmission of AAFVs). We extracted estimates and 95% CI for the average response of each mosquito genera–viral group pairing from the fitted models using the function *emmeans* in the package *emmeans* [39]. Finally, we estimated the strength of the overall correlation between the maximum detected proportions of mosquitoes infected and transmitting across all mosquito–virus pairs using a simple correlation test (using the R function *cor*.*test*).

For these statistical analyses we focused on infection and transmission because they are the critical components of vector competence and were historically measured most frequently in experiments. Quantifying dissemination of the virus is a relatively recent addition to measurements used to assess the vector competence of mosquitoes for arboviruses [18], so including this metric in our statistical testing may limit the inclusion of studies reported prior to its introduction.

#### Dose- and time-dependent measures

Continuous dose- and time-response curves can better capture the full spectrum of conditions experienced by mosquitoes feeding on viremic vertebrate hosts in real-world conditions. However, a lack of comprehensive data for all mosquito-virus pairings prevented us from fully examining dose- and time-dependent infection, dissemination, and transmission for each mosquito-virus pair. As sufficient data was available, we fitted time- and dose-dependent regression models for *Ae. aegypti* as a case study to: 1) explore variation in the shapes of infection and transmission curves across viruses; and 2) highlight the differences between a single metric obtained across all experiments (e.g., maximum observed) and the full infection and transmission functions estimated using all available experimental data. We fitted the proportion of *Ae. aegypti* infected, disseminated, and transmitting using generalized linear mixed effects models (GLMM) with Binomial error distributions in R using the package *lme4* [40]. For each model, the proportion of mosquitoes that became infected, developed a disseminated infection, or transmitted the virus was taken as the response variable and the total numbers of mosquitoes exposed were used as weights.

The model for infection included an interaction between virus species and virus dose, a fixed effect for DPE, and a random intercept of “study” to control for variation among citations not captured by the included fixed effects. The models for both dissemination and transmission included an interaction between virus species and DPE, a fixed effect for virus dose, and a random intercept for citation. Logistic regression assumes that probability reaches an asymptote of one, which implies that at some dose and day, all *Ae. aegypti* individuals are expected to become infected, develop a disseminated infection and transmit each virus. While a model that explicitly estimates the asymptotic value is desirable, we were unable to estimate variable asymptotes by virus species due to paucity of data, even for the relatively well-studied *Ae. aegypti*. Confidence intervals for the fitted models were obtained using 500 parametric bootstraps.

## Results

### Virus group and mosquito genera associations

#### Aedes*-associated flaviviruses*

Only two Australian mosquito genera, *Aedes* and *Culex*, have been tested for their competence for AAFVs. Of these, a significantly larger proportion of *Aedes* individuals became infected with (mean: 43%; 95% CI: 39%-48%) and could transmit (mean: 30%; 95% CI: 25%-36%) these viruses than *Culex* individuals (infection: mean: 4%; 95% CI: 1.1%-13%; transmission: mean: 0.9%; 95% CI: .05%-13%) (Fig 2).

**Figure 2.**
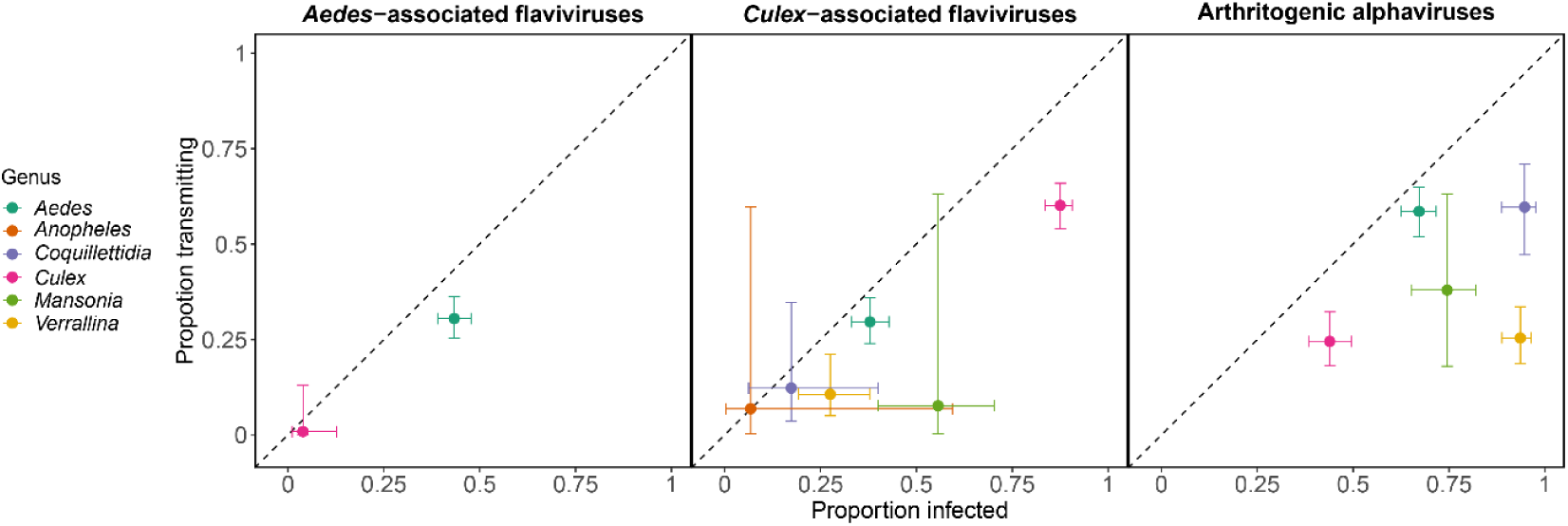
Estimated proportion of mosquitoes (grouped by genus: color) infected by (x-axis) and transmitting (y-axis) three virus groups (panels). Points show mean estimates and intervals show 95% confidence intervals (CI) obtained from the logistic regression models. Non-overlapping horizontal and vertical CI between two mosquito genera for a given viral group indicate statistically different infection and transmission capabilities, respectively, for those genera.

#### Culex*-associated flaviviruses*

A significantly higher proportion of *Culex* mosquitoes became infected with CAFVs (mean: 88%, 95% CI: 84%-91%) than any of the other five genera; significantly more *Culex* transmitted (mean: 60%, 95% CI: 54%-66%) than *Aedes, Coquillettidia*, and *Verrallina* (Fig 2). Wide 95% confidence intervals for both *Anopheles* and *Mansonia*, due to small sample sizes in these genera (8-9 mosquitoes tested), obscured our ability to statistically differentiate their transmission ability from that of *Culex* despite large differences in their estimated means (Fig 2). The CAFVs were the only group in the data set that contained at least one response from each mosquito genus for both infection and transmission, which strengthens the evidence for the importance of *Culex* as vectors of these viruses.

#### Arthritogenic alphaviruses

There were some clear differences among mosquito genera for vector competence for arthritogenic alphaviruses. In particular, the largest proportion of *Coquillettidia* and *Verrallina* mosquitoes became infected with arthritogenic alphaviruses, although more *Coquillettidia* transmitted these viruses than *Verrallina* (Fig 2). Although *Aedes* were generally less susceptible to infection than *Coquillettidia*, a similar proportion transmitted the virus (Fig 2). Overall, arthritogenic alphaviruses stood out as having the strongest ability to infect and be transmitted by multiple mosquito genera. A larger proportion of mosquitoes across all genera became infected with and transmitted arthritogenic alphaviruses than for CAFVs (difference in infection: β =1.21, SE = 0.17, p < 0.05; transmission: β =1.25, SE = 0.20, p < 0.05; coefficient on the logit-scale) and AAFVs (difference in infection: β = 0.98, SE = 0.15, p < 0.05; transmission: β = 1.21, SE = 0.19, p < 0.05; coefficient on the logit-scale).

### Variation within virus groups and mosquito genera

These results reveal many clear differences in the mean response of mosquito genera to exposure to different virus groups (Fig 2). However, mosquito species within a genus vary substantially in their ability to became infected with, disseminate, and transmit each of the virus species within each viral group.

#### Aedes*-associated flaviviruses*

*Aedes* spp. varied strongly in their ability to become infected with different AAFVs, from a minimum of 3% of *Aedes notoscriptus* individuals becoming infected with DENV-4 to a maximum of 100% of *Ae. aegypti* individuals becoming infected with each DENV strain and 100% of *Aedes albopictus* individuals becoming infected with ZIKV (Fig 3B). Similar to the variation in infection, *Aedes* species also varied in their ability to transmit AAFVs, from zero percent of *Aedes katherinensis* transmitting DENV-2, or *Aedes procax* and *Aedes vigilax* transmitting ZIKV, to a maximum of 87% of *Ae. aegypti* transmitting ZIKV (Fig 4B). In contrast to the large variation among *Aedes* spp., none of the three *Culex* species (*Culex annulirostris, Culex quinquefasciatus*, and *Culex sitiens*) experimentally exposed to ZIKV were able to transmit the virus (Fig 4B) and only one individual *Cx. quinquefasciatus* (of 50 tested) was infected (Fig 3B). Similarly, *Cx. annulirostris* was refractory to infection with DENV-4 in the only experiment where Australian *Culex* spp. had been exposed to any of the DENVs (Fig 3B).

**Figure 3.**
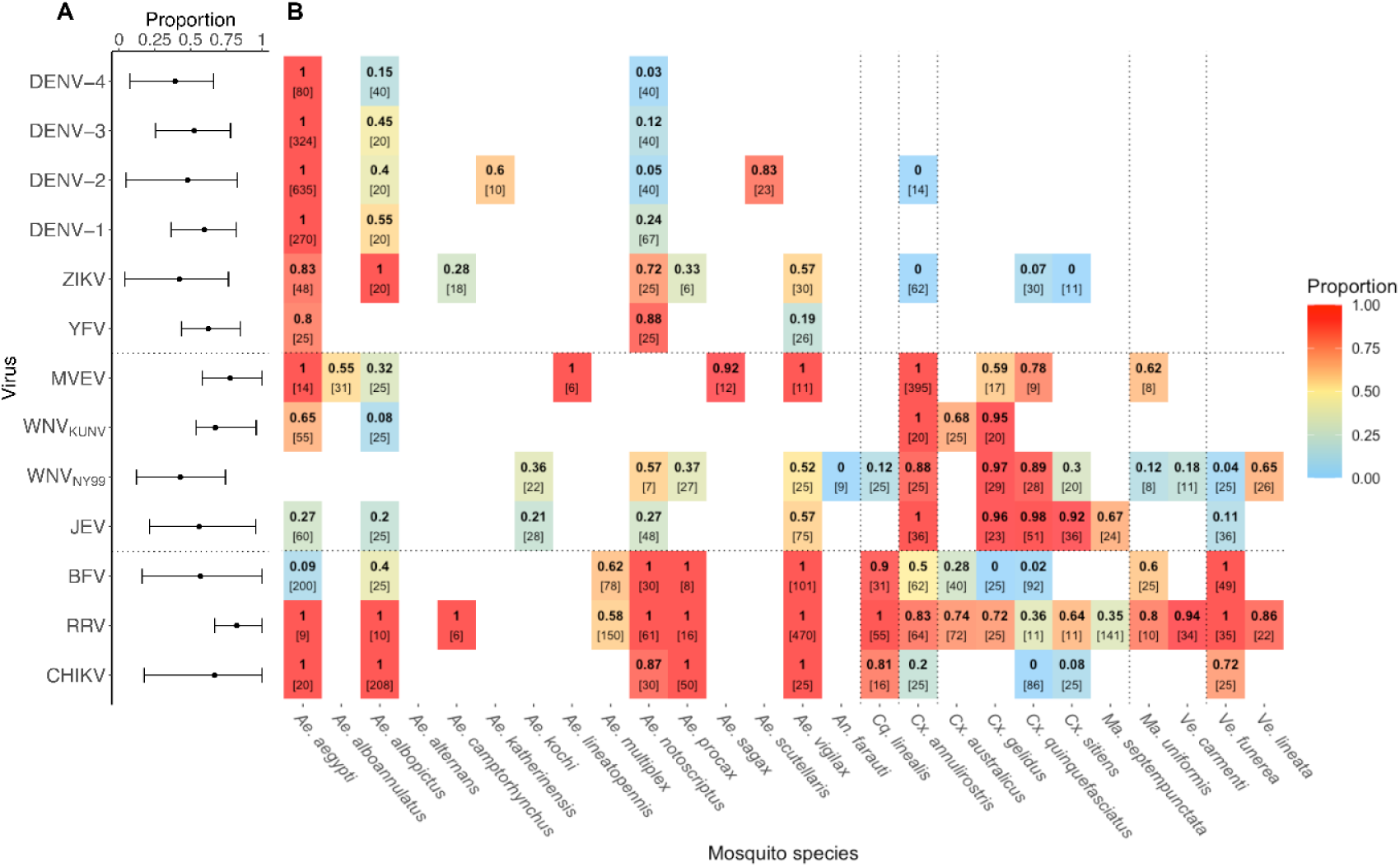
**A)** Maximum proportion of mosquitoes positive for infection across all mosquito species tested for each virus (median and 80% quantiles across all mosquitoes). **B)** Maximum proportion of each mosquito species found positive for infection among all of the laboratory experiments for each mosquito-virus pair (represented by color and the bold-face top number in each box). The number of experimental mosquitoes tested in the study[s] in which the maximum proportion was observed is displayed in square brackets (in the case of multiple experiments finding the same maximum, the sample sizes were summed). Viruses are organized into three groups: *Aedes-associated flaviviruses* (yellow fever [YFV], Zika [ZIKV] and dengue [DENV, separated by serotype]); *Culex-associated flaviviruses* (Japanese Encephalitis [JEV], West Nile New York strain [WNV_NY99_], West Nile Kunjin strain [WNV_KUNV_] and Murray Valley encephalitis [MVEV]); and *arthritogenic alphaviruses* (chikungunya [CHIKV], Ross River [RRV] and Barmah Forest [BFV]). Vertical dotted lines separate mosquito genera, from left to right: *Aedes* [*Ae*], *Anopheles* [*An*], *Coquillettidia* [*Cq*], *Culex* [*Cx*], *Mansonia* [*Ma*] and *Verrallina* [*Ve*].

**Figure 4.**
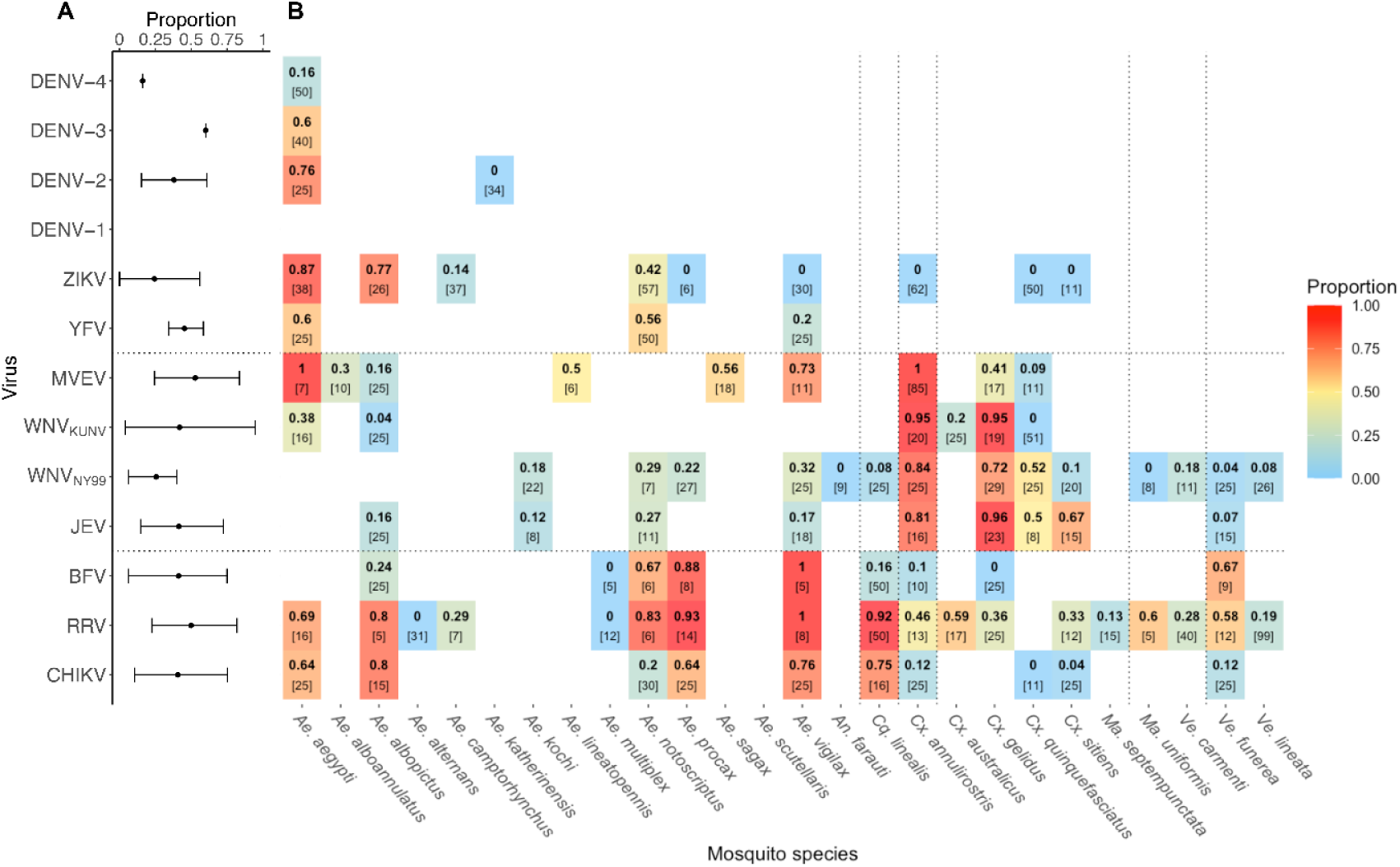
**A)** Maximum proportion of mosquitoes positive for transmission across all mosquito species tested for each virus (median and 80% quantiles across all mosquitoes). **B)** Maximum proportion of each mosquito species found positive for transmission among all of the laboratory experiments for each mosquito-virus pair (represented by color and the bold-face top number in each box). The number of experimental mosquitoes tested in the study[s] in which the maximum proportion was observed is displayed in square brackets (in the case of multiple experiments finding the same maximum, the sample sizes were summed). Viruses are organized into three groups and mosquito species are grouped into genera as in Figure 2.

#### Culex*-associated flaviviruses*

Variation among the CAFVs was more difficult to generalize than that of the other viral groups. MVEV had the highest average overall infection and transmission proportions (78% and 53%, respectively) of all viruses within the group, and one of the highest across all viruses (Fig 3, 4). By contrast, WNV (New York 1999 strain; WNV_NY99_) had the lowest infection and transmission proportions (43% and 26%, respectively) of all CAFVs (Fig 3, 4). All *Culex* species were highly susceptible to infection with CAFVs except *Cx. sitiens* exposed to WNV_NY99_ (Fig 3B). *Cx. annulirostris* and *Culex gelidus* transmitted the CAFVs at rates above 40% (Fig 4B). Although not as susceptible as *Culex* species for the CAFVs, the majority of *Aedes* spp. including *Ae. aegypti, Ae. vigilax, Aedes sagax*, and *Aedes lineatopennis* demonstrated relatively high infection proportions (>50%) with MVEV (Fig 3B). All examined *Verrallina* and *Mansonia* species were susceptible to infection with CAFVs, but in almost all cases, the proportion infected and transmitting these viruses were lower than *Culex* spp. (Fig 3B, 4B).

#### Arthritogenic alphaviruses

The high susceptibility to infection and subsequent transmission of arthritogenic alphaviruses by most Australian mosquitoes (Fig 3, 4) potentially points to a general virus genus attribute, especially given the high number of mosquito species (19 out of 27) representing different genera that were exposed to these viruses. *Aedes* spp. were generally highly susceptible to infection and readily transmitted most arthritogenic arboviruses (Fig 3, 4). For example, both *Ae. aegypti* and *Ae. albopictus* were highly susceptible to RRV and CHIKV, and readily transmitted these viruses at rates above 60%. However, both species had low infection proportions for BFV (< 9% and ≤ 40%, respectively; Fig 3B). There was more variation among *Culex* spp. in competence for arthritogenic alphaviruses than for other mosquito genera. All species of *Culex* were highly susceptible to infection with RRV (> 60%) but had very low susceptibility to CHIKV and BFV (Fig 3B). All *Culex* spp. were capable of transmitting RRV, although they had a much lower probability than either *Aedes* or *Coquillettidia. Verrallina* were highly susceptible to infection with all anthritogenic alphaviruses, but like *Culex*, had lower transmission proportions than other mosquito genera tested (Fig 3, 4).

### Correlations among Infection, Dissemination, and Transmission

Across all mosquito species and viruses, we found a strong positive correlation (r = 0.80, 95% CI: 0.71-0.87) between mosquito species’ maxima for infection and transmission (Fig 5). While some of this correlation can be attributed to research effort (mosquitoes studied more for infection are also studied more for transmission (S1 Fig), and detected maxima increase with increased sample sizes (p < 0.05, S2 Fig)), the strength of this correlation points to the strong biological link between infection and transmission within mosquitoes.

**Figure 5.**
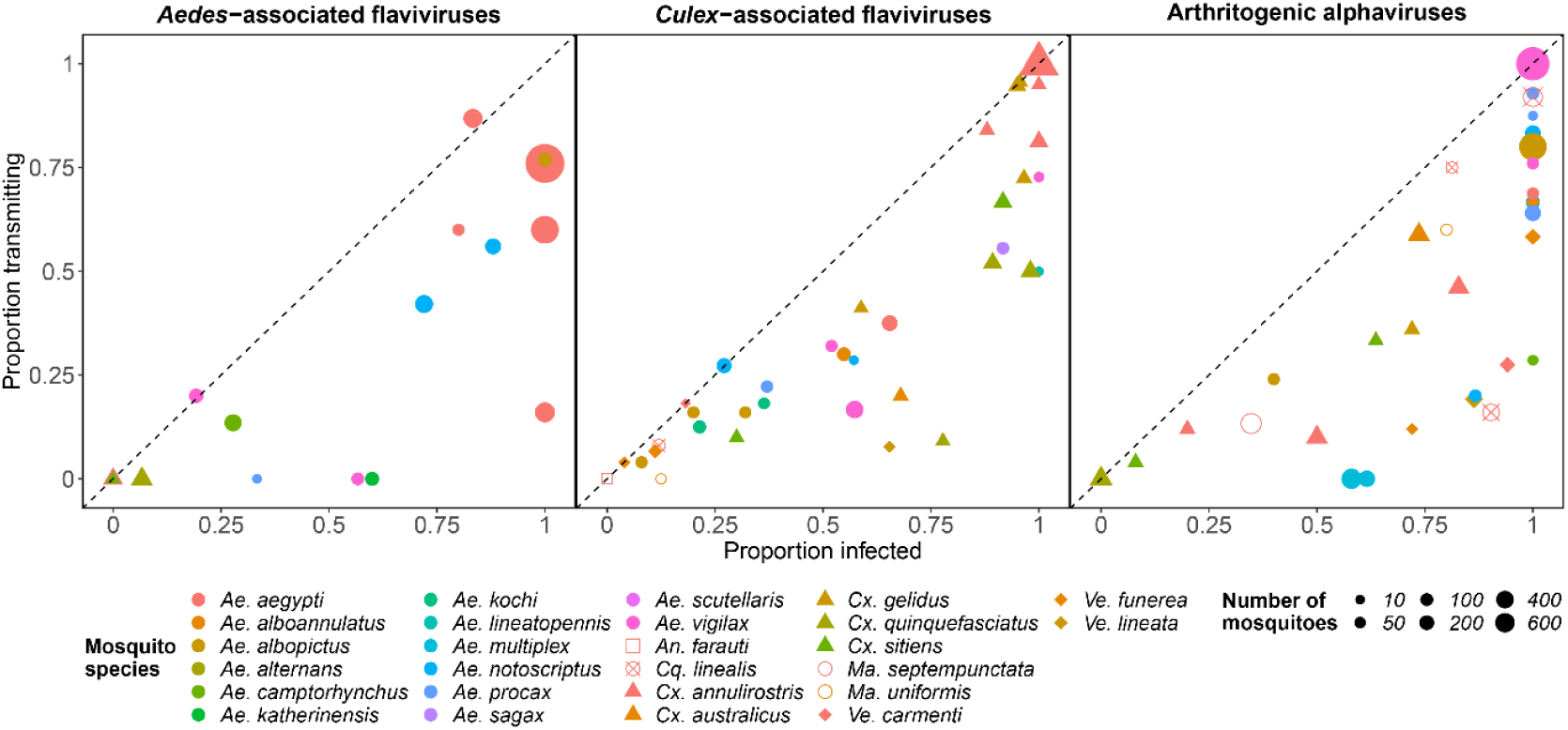
Infection and transmission probabilities for each mosquito-virus pairing tested. Multiple points of the same color and shape within a viral group represent results from different viruses within that group. The dashed line is a 1:1 line; points beneath the line represent mosquito-virus pairings with a greater infection probability than transmission probability.

*Aedes* spp. typified this relationship, falling close to the 1:1 line, with similar infection and transmission probabilities for all viral groups (Fig 5). Other mosquito genera and many individual species stood apart from this general trend. For instance, *Verrallina lineata* had the largest difference between their average (across all species) maximum infection and transmission probabilities (infection: 76%; transmission: 13%) for the two viruses (WNV and RRV) they were exposed to in our dataset. Similarly, those *Culex* spp. that did become infected with arthritogenic alphaviruses transmitted these viruses at very low rates, especially when compared with other genera (Fig 3, 4). Among all mosquitoes tested for both infection and transmission with at least five viruses, *Cx. quinquefasciatus* had the largest difference (infection: 53%; transmission: 19%) and *Ae. notoscriptus* had the smallest difference in average observed maxima across all viruses (infection: 52%; transmission: 49%).

While these data make it difficult to quantitatively compare the impacts of each barrier (only 19% of all mosquito-virus pairings were assessed for infection, dissemination, and transmission, and only a small subset of these were assessed in the same laboratory setting), we obtained a coarse measure of the proportion of mosquitoes where the virus was unable to pass through each successive barrier using a sample size weighted average across all mosquito-virus pairs for each barrier. Across all mosquito-virus pairs and all experiments, conditional on exposure, 72% of mosquitoes became infected, 56% of mosquitoes disseminated virus, and 38% transmitted, implying that on average 72% of mosquitoes became infected, 77% of infected mosquitoes developed a disseminated infection, and 68% of mosquitoes with a disseminated infection went on to transmit. Although this method provides only a coarse measure of barrier importance relative to using single studies measuring infection, dissemination, and transmission for each vector – virus pair [41], these results indicate that no single barrier stands out as the sole restriction to transmission ability, suggesting that all three are important.

### Variation among experimental design and study effort

Vector competence studies vary greatly in their experimental design, which can influence study outcomes. In particular, the experimental doses that mosquitoes were exposed to varied strongly both within and between viruses (S3 Fig, S4 Fig). For example, only a single dose was used for WNV_NY99_ (6.7 log_10_ infectious units (IU)/mL)), while doses spanned a wide range for other viruses, capturing more of the potential range of natural exposure to a viremic host (e.g., 3.2 to 10 log_10_ IU/mL for DENV-4 and 2.9 to 9.3 log_10_ IU/mL for RRV; S3 Fig). For species that were exposed to a range of virus doses, the dose that infected the maximum number of mosquitoes was generally among the higher of the doses that the mosquitoes were exposed to (S3 Fig). Similarly, the DPE on which mosquitoes were tested also varied among studies (S3 Fig - S6 Fig).

### *Dose- and time-dependent vector competence responses of* Aedes aegypti *exposed to dengue, chikungunya and Zika viruses*

As a case study, we used a GLMM to model the continuous dose- and time-dependent responses (instead of just maximum potential) of *Ae. aegypti*, which has been the subject of the most extensive assessment of vector competence of all Australian mosquito species. Even for the best-studied pairings, *Ae. aegypti* exposed to CHIKV, ZIKV, and DENV-3, confidence intervals were wide mostly due to few data points and highly variable results at single doses and days across experiments (S7 Fig). Despite this uncertainty, these dose- and time-dependent continuous functions help to illustrate how relying on a single maximal competence measure could bias estimates of competence in practice, as it does not capture the conditions mosquitoes will experience when transmitting under natural conditions. However, until the substantial uncertainty is reduced, estimates from dose- and time-dependent continuous functions are unlikely to be more useful than a maximal measure.

## Discussion

Our analysis of vector competence experiments, involving 115 mosquito-virus pairings across 295 total experimental treatments, quantified virus group–mosquito genus associations, corroborated existing canonical groupings, and refined our understanding of variation in the ability for different mosquito species to become infected with and transmit different arboviruses. Our synthesis reinforced the canonical groupings of *Aedes-* and *Culex-* associated flaviviruses; *Aedes* spp. were more competent than *Culex* spp. for AAFVs (the only two genera tested in our data set), whilst *Culex* spp. had the highest infection and transmission potential for CAFVs of all six genera tested (Fig 2). The association of CAFVs with *Culex* spp. in the Australian context is supported by field studies that have detected the majority of CAFVs in *Culex* spp., particularly *Cx. annulirostris* [42-44]. Though AAFVs are not endemic to Australia, a few instances of field evidence of AAFV carriage by Australian mosquitoes provide some independent support for our vector competence-based *Aedes-* flaviviruses associations. For example, DENV-2 RNA was detected in *Ae. aegypti* collected during a dengue outbreak in north Queensland in 2003 [45], and early experiments by Bancroft [46] and Cleland et al. [47] conducted during dengue outbreaks in the early 20^th^ century in Australia demonstrated that *Ae. aegypti* were infected with DENV as evidenced by transmission of the viruses to susceptible humans. Further, no *Cx. quinquefasciatus* tested by Cleland et al. [47] transmitted the virus, suggesting a low or negligible field infection rate and reinforcing the overall poor vector competence of members of the *Culex* genus for AAFVs. There have been no reported autochthonous cases of ZIKV or YFV in Australia.

In contrast to the flaviviruses, our analysis revealed that no single mosquito genus was clearly the most competent for infection with and transmission of the alphaviruses, which fits their previous description of vector promiscuity [48, 49]. Broadly, *Culex* spp. had the lowest vector competence, *Coquillettidia* had the highest vector competence, and *Aedes* spp. had average susceptibility to infection but higher than average ability to transmit alphaviruses (Fig 2, 5). These results concur with the plasticity in vector utilization observed for the arthritogenic alphaviruses [49] and the diversity of mosquitoes found infected with RRV and BFV in Australian mosquito populations [50-52]. Like the exotic flaviviruses, to date, there has been no evidence of local transmission of CHIKV nor field isolation of this virus from Australian mosquitoes, despite the presence of populations of highly competent vectors, including *Ae. aegypti* and *Ae. albopictus* in the state of Queensland and close geographical proximity of Australia to CHIKV endemic regions, such as Indonesia.

As vector competence tends to cluster by mosquito genus and virus sub-family (Fig 2), using genus and family-level information could help to constrain uncertainty about an unmeasured mosquito species-virus pair. This is potentially useful for predicting the consequences of an invading virus into a location with indigenous mosquito fauna not historically associated with the virus, or for the range expansion of a mosquito species that brings them into contact with circulating viruses in their new ecological niche. Whilst mosquito genus-level virus group associations were relatively robust, we did find substantial variation in vector competence at the mosquito species level (Fig 3-5). Although many of the *Aedes* spp. established as global vectors of AAFVs had high competence for many AAFVs, we observed substantial variation in competence among *Aedes* species for individual AAFVs (Fig 3, 4). Commensurate with its role globally, Australian *Ae. aegypti* are highly susceptible to infection and readily transmit DENV, YFV and ZIKV. In contrast, *Ae. notoscriptus*, which shares an overlapping ecological niche with *Ae. aegypti*, was an efficient laboratory vector of YFV, varied in response to challenge with different ZIKV strains, and was a poor vector of DENVs (Fig 3, 4). Although it could not be captured in our analysis, there is also considerable intraspecific variation in vector competence. For instance, *Cx. annulirostris* and *Ae. aegypti* collected from different locations in Australia significantly differed in their response to exposure to WNV [53, 54] and DENVs [55], respectively.

There are numerous intrinsic and interacting viral and vector traits that could determine the specificity or promiscuity of arbovirus-mosquito associations. Amino acid changes, even as a result of a single point mutation in an envelope protein, can facilitate virus adaptation to local mosquito vectors increasing the efficiency of their transmission [56-58]. While mutations that give rise to virus adaptation have the capacity to occur through a single passage in the mosquito [59], they are more likely to arise after many transmission generations. We found multiple instances where viruses were efficiently transmitted by a mosquito species they have never encountered in the field, a situation exemplified by the high vector competence of *Ae. vigilax* and *Ae. procax* for CHIKV, and of *Ae. notoscriptus* for YFV. This indicates that viral mutations and adaptation to local mosquito species are not necessarily a prerequisite for transmission by mosquito species the virus has not encountered. This supports a study in Thailand that found no evidence of adaptation of sympatric or allopatric isolates of DENVs to local *Ae. aegypti* populations [60]. It also indicates that some molecular determinants of vector competence may be conserved within mosquito and/or virus taxa, while others may be more specific. For example, *Aedes* spp. potentially possess a receptor or multiple receptors on the mesenteronal epithelial cells to which the envelope glycoproteins on the surface of AAFVs bind, thus initiating infection of the mesenteron [12]. In contrast, *Culex* spp. may not possess these specific receptors (or they may be restricted in density or diversity), limiting the number of binding sites for this group of viruses. Other intrinsic factors that influence virus–vector interactions are linked to diversity in the mosquito microbiome [61] and virome [62], and antiviral mechanisms in the mosquito [63]. Any of these factors could not only explain the virus-mosquito associations described above, but also how viruses that invade a virgin ecosystem are able to exploit the resident mosquito population, as occurred with WNV in North America [11] and establishment of YFV in *Haemagogus*/*Sabethes*-driven sylvatic cycles in South America [64].

We also found considerable variation in experimental conditions among the vector competence studies; mosquito and virus genotype, temperature, virus dose (S3 Fig), the DPE on which mosquitoes were tested (S4 Fig - S6 Fig), and the nutritional status of the mosquito are just a few of the many factors that varied among studies that have been known to strongly influence the outcomes of individual mosquito-virus pairings [14, 22]. Further, experimental methods, such as the feeding protocol used to expose mosquitoes to the virus (e.g., live animal, membrane or blood-soaked pledget), virus isolate and passage used, origin of mosquito, mosquito laboratory generation number, and method used to assess transmission or assay to detect the presence of virus varied greatly among studies (see Online Supplemental Data Files).

On one hand, such variation makes it more difficult to predict the realized ability of a given mosquito to transmit a given virus and likely added much experimental uncertainty to the intrinsic variation within mosquito genera-virus family groupings. On the other hand, identifying the conditions a species requires to reach maximal transmission potential requires variation between and within experiments given the vast amount of variation that occurs during a transmission cycle under natural conditions. That is, when mosquitoes have been infected with only a single dose (e.g., see WNV_NY99_ in S3 Fig), and measured at the same time points (e.g., see YFV in S4 Fig), we are limited to an understanding of which species are more competent under a narrowly-bounded setting and not the range of conditions that a mosquito would experience in nature when biting viremic hosts. By contrast, when mosquitoes are exposed to a wide range of doses and measured on many different DPE (e.g., see RRV in S3 Fig - S6 Fig), we are better able to capture variation in maxima among species and can take steps to quantify the full dose- and time-dependency of infection and transmission.

Dose-and time-dependent infection and transmission functions that utilize the full range of experimental conditions are powerful tools for quantifying the role different mosquito species play in viral transmission cycles. Specifically, when these continuous functions are combined with additional data, such as host viremia, mosquito feeding patterns, and/or climate conditions favorable to mosquito longevity, we are best able to gain understanding of the circumstances that allow different species to have increased transmission potential (i.e., vectorial capacity; see [65, 66]). Regression models like those we used for *Ae. aegypti* (S7 Fig) can provide the required data but are useful for only very well studied mosquito-virus pairs as they are more data intensive than calculating a maxima. Ultimately, using the maxima is necessary for most mosquito-virus pairs (for many of which there is little data) and despite collapsing much variation it enables rapid incrimination of mosquitoes as arbovirus vectors.

This synthesis of vector competence studies provides a foundational tool for understanding the vector competence of mosquitoes for many of the medically important arboviruses. Although we focused our analysis on mosquitoes originating from Australia, a synthesis of vector competence experiments from around the world would potentially reinforce and expand on the outcomes of the current study. The degree of variation within each of the taxonomic levels highlights that although general virus–mosquito associations can help incriminate mosquitoes as vectors of arboviruses, location-specific experiments assessing the susceptibility and transmission ability of uncharacterized virus–mosquito pairing are still critical for establishing the role of a mosquito species in transmission cycles. When combined with data on vector abundance and human contact patterns [67], this information can guide public health responses to arbovirus outbreaks with view to limiting the disease incidence in susceptible populations.

## Data Availability

All data produced in the present work are uploaded in a supplemental file

## Acknowledgements

We thank Marissa Childs and Nicole Nova for their feedback on early versions of the analysis and data visualizations. We also thank John Mackenzie and Alyssa Pyke for discussions on virus phylogeny and evolution. Alyssa Pyke also provided valuable comments on the manuscript. EAM was supported by the National Science Foundation and the Fogarty International Center (DEB-2011147), Stanford’s Center for Innovation in Global Health, Woods Institute for the Environment, and King Center for Global Development, and the Terman Award. EAM, MPK, and EBS were supported by the National Institute of General Medical Sciences (R35GM133439). MPK was supported by the Natural Capital Project. AvdH states that: ‘‘The opinions, interpretations and conclusions are those of the author and do not necessarily represent those of the organization.’’

## Data availability

All data and code used in this study are available in the online supplemental materials. Code is hosted at: https://github.com/morgankain/VectorCompetence

## Supporting information

**S1 Fig. Number of mosquitoes of each species assessed for infection and transmission in vector competence experiments**. Mosquito species frequently measured in infection experiments (total sample size across all experiments; X axis) were also more frequently measured in transmission experiments (total sample size across all experiments; Y axis). Each point for a mosquito species represents a different virus.

**S2 Fig. Total number of mosquitoes assessed for infection, dissemination and transmission in vector competence experiments**. Increased research effort leads to a higher maximum proportion for infection, dissemination, and transmission (logistic regression, p < 0.05). Increased dose does not lead to a detectable increase in maximum proportion (logistic regression, p > 0.05). Each point represents a single virus-species pair.

**S3 Fig. Viral doses mosquitoes were exposed to across all laboratory experiments that measured infection**. Blue points show experimental doses that did not lead to the highest detected proportion of infected mosquitoes; red points show the infectious dose[s] that resulted in the highest observed infected proportion.

**S4 Fig. Days post viral exposure on which mosquitoes were tested for infection status**. Blue points show experimental days that did not lead to the highest detected proportion of infected mosquitoes; red points show the day[s] that resulted in the highest observed infected proportion.

**S5 Fig. Viral doses mosquitoes were exposed to across all laboratory experiments that measured transmission**. Blue points show experimental doses that did not lead to the highest detected proportion of transmitting mosquitoes; red points show the infectious dose[s] that resulted in the highest observed transmitting proportion.

**S6 Fig. Days post viral exposure on which mosquitoes were tested for their ability to transmit viruses**. Blue points show experimental days that did not lead to the highest detected proportion of transmitting mosquitoes; red points show the day[s] that resulted in the highest observed transmitting proportion.

**S7 Fig. Continuous functions for mosquito infection over dose (top), and for dissemination and transmission over time (middle and bottom, respectively)**. The results pictured here are from a generalized linear mixed effects model (GLMM) using *Aedes aegpyti* for all viruses they have been tested with in the lab; however, here we only show the three viruses for which more than one data point exist for infection, dissemination, and transmission. For infection (top), medians (solid lines) and 95% confidence intervals (grey bands) are estimated using twelve days post inoculation; for dissemination (middle) and transmission (bottom) estimates are drawn for a dose of 7 log10 IU/mL.

**S1 File. Australian mosquito vector competence source data**.

