## Supplemental Figures for "Not all mosquitoes are created equal: incriminating mosquitoes as vectors of arboviruses"

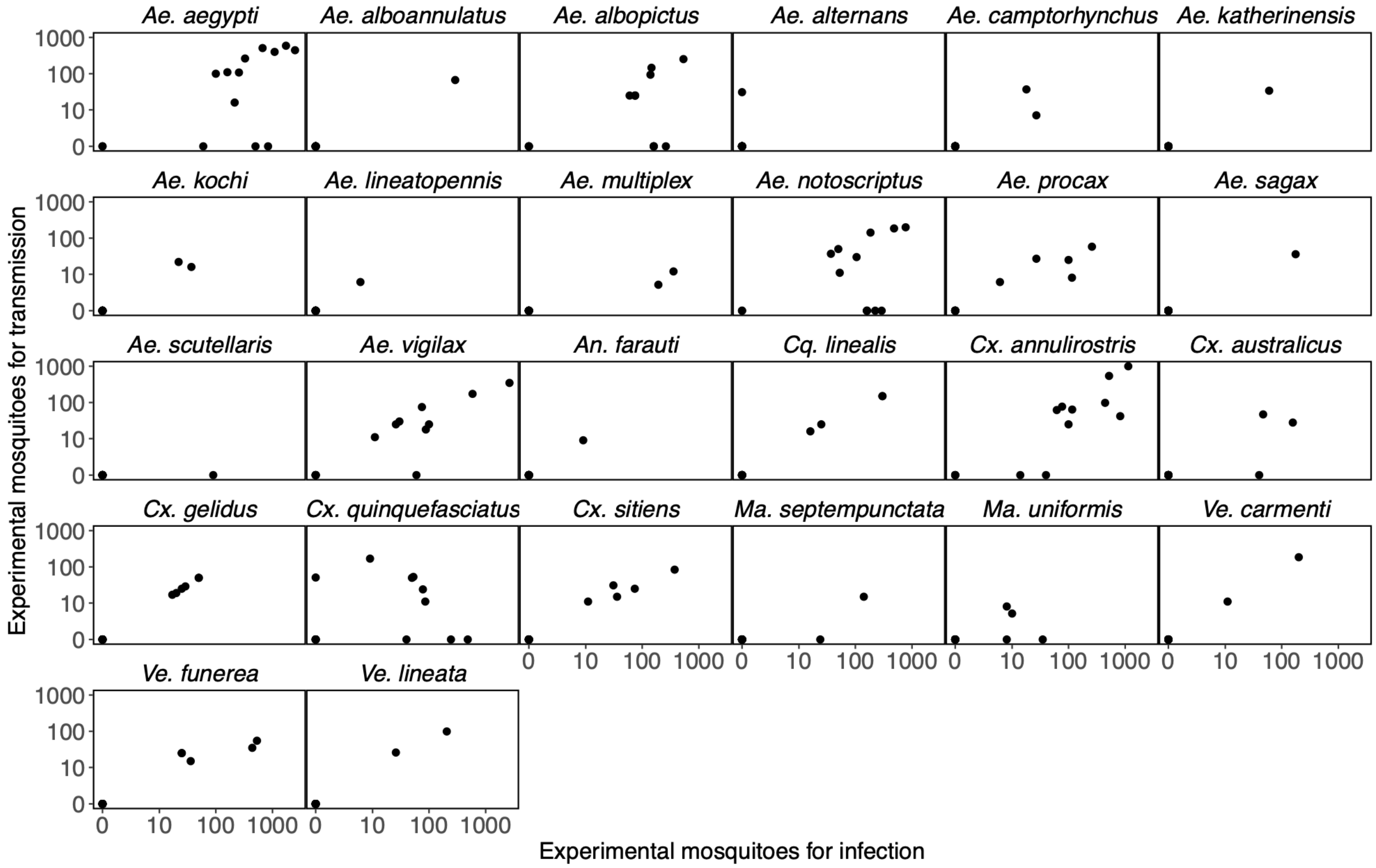


**S1 Fig**. **Number of mosquitoes of each species assessed for infection and transmission in vector competence experiments.** Mosquito species frequently measured in infection experiments (total sample size across all experiments; X axis) were also more frequently measured in transmission experiments (total sample size across all experiments; Y axis). Each point for a mosquito species represents a different virus.

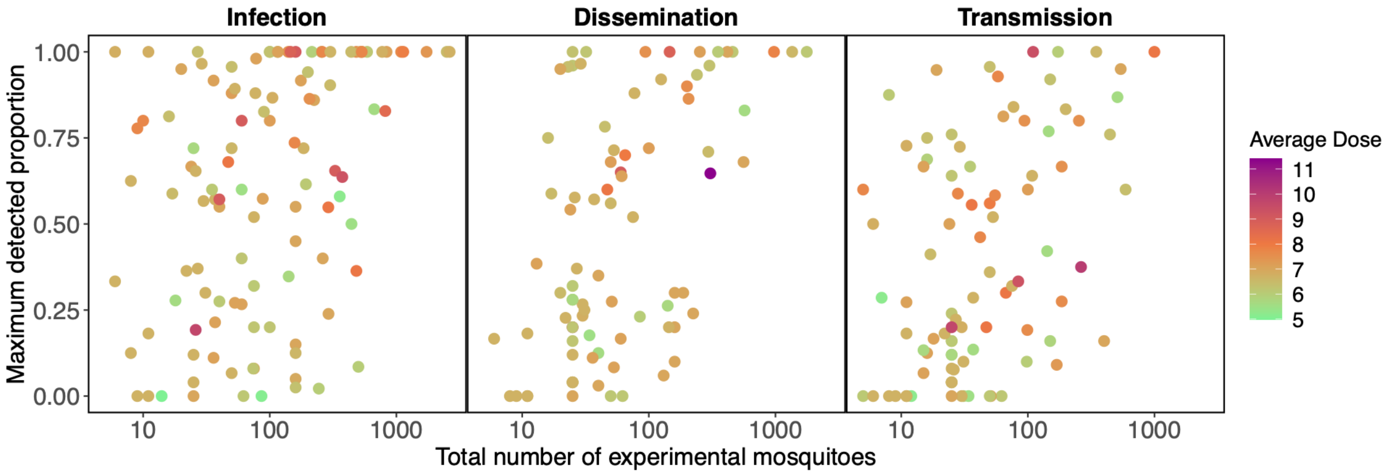


**S2 Fig**. **Total number of mosquitoes assessed for infection, dissemination and transmission in vector competence experiments.** Increased research effort leads to a higher maximum proportion for infection, dissemination, and transmission (logistic regression, $p < 0.05$). Increased dose does not lead to a detectable increase in maximum proportion (logistic regression, $p > 0.05$). Each point represents a single virus-species pair.

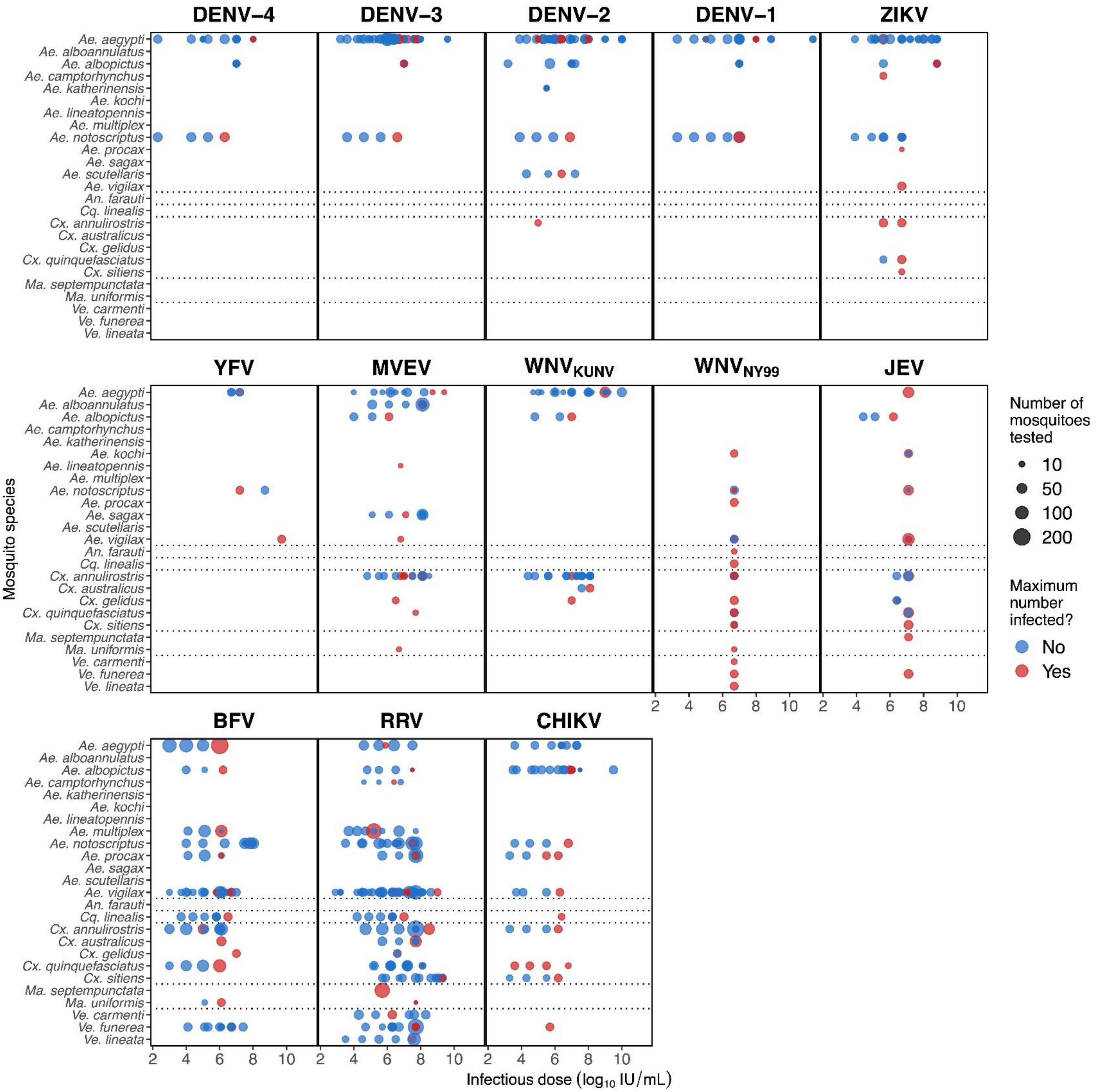


**S3 Fig**. **Viral doses mosquitoes were exposed to across all laboratory experiments that measured infection.** Blue points show experimental doses that did not lead to the highest detected proportion of infected mosquitoes; red points show the infectious dose[s] that resulted in the highest observed infected proportion.

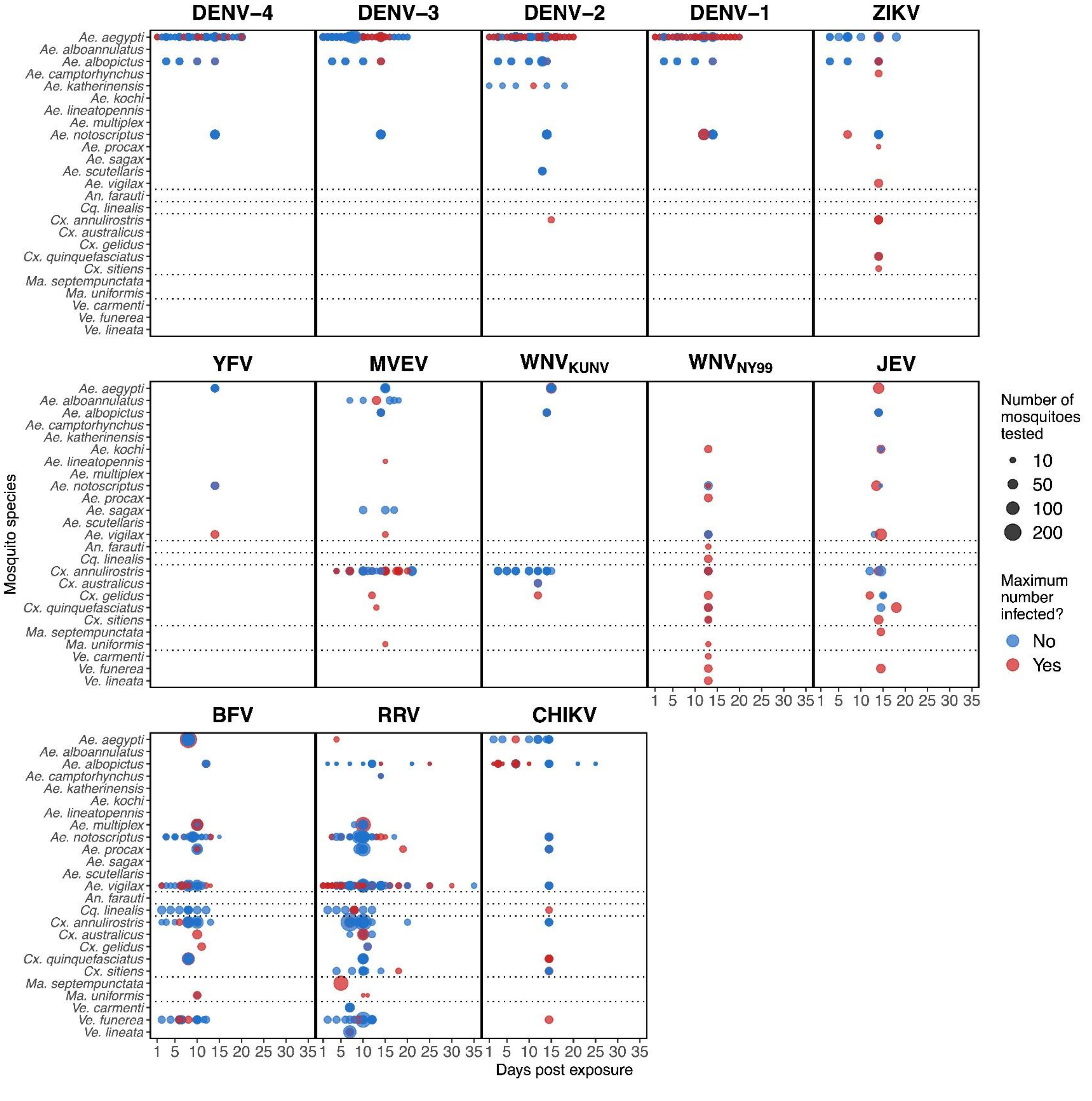


**S4 Fig**. **Days post viral exposure on which mosquitoes were tested for infection status.** Blue points show experimental days that did not lead to the highest detected proportion of infected mosquitoes; red points show the day[s] that resulted in the highest observed infected proportion.

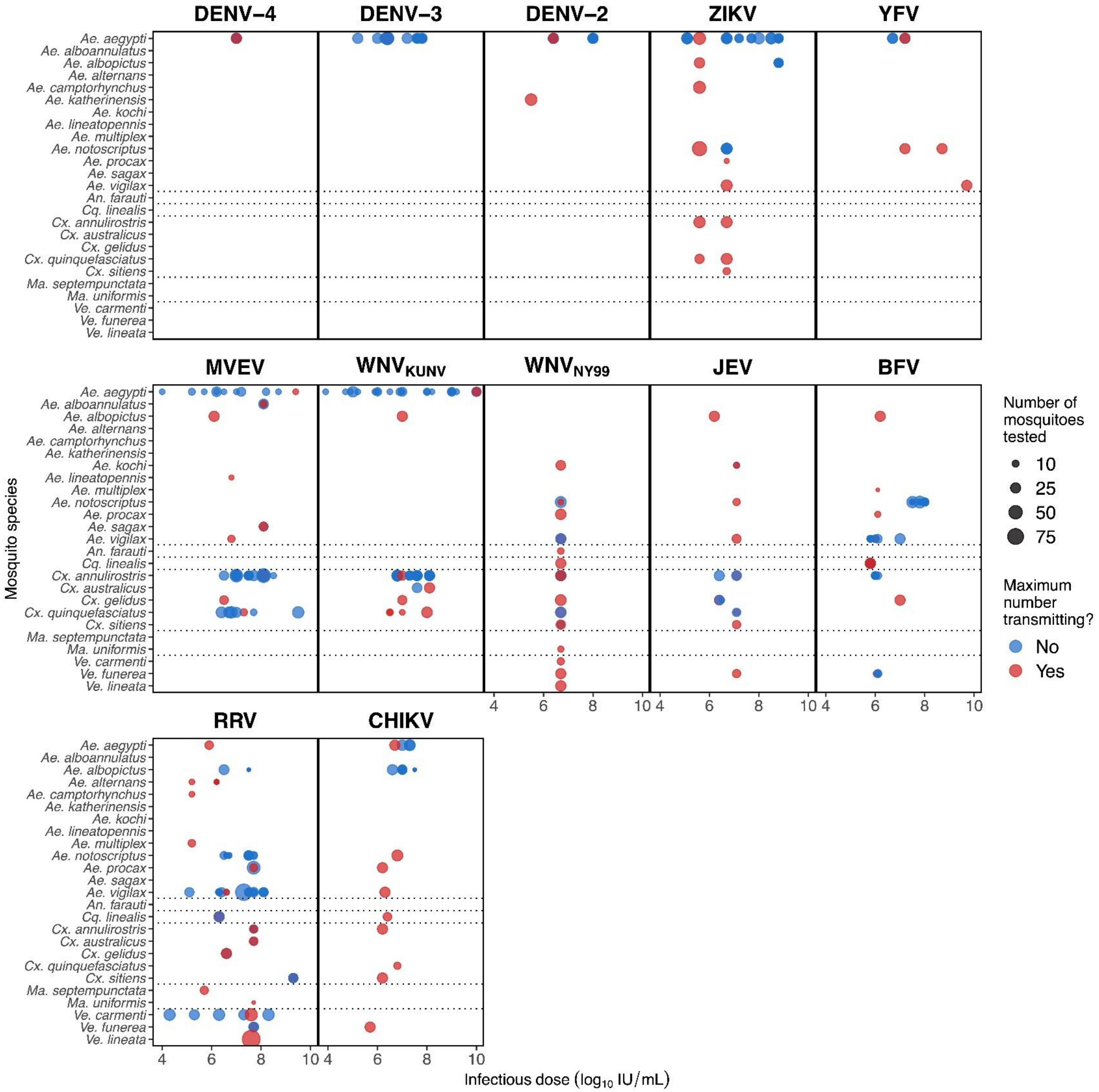


**S5 Fig**. **Viral doses mosquitoes were exposed to across all laboratory experiments that measured transmission.** Blue points show experimental doses that did not lead to the highest detected proportion of transmitting mosquitoes; red points show the infectious dose[s] that resulted in the highest observed transmitting proportion.

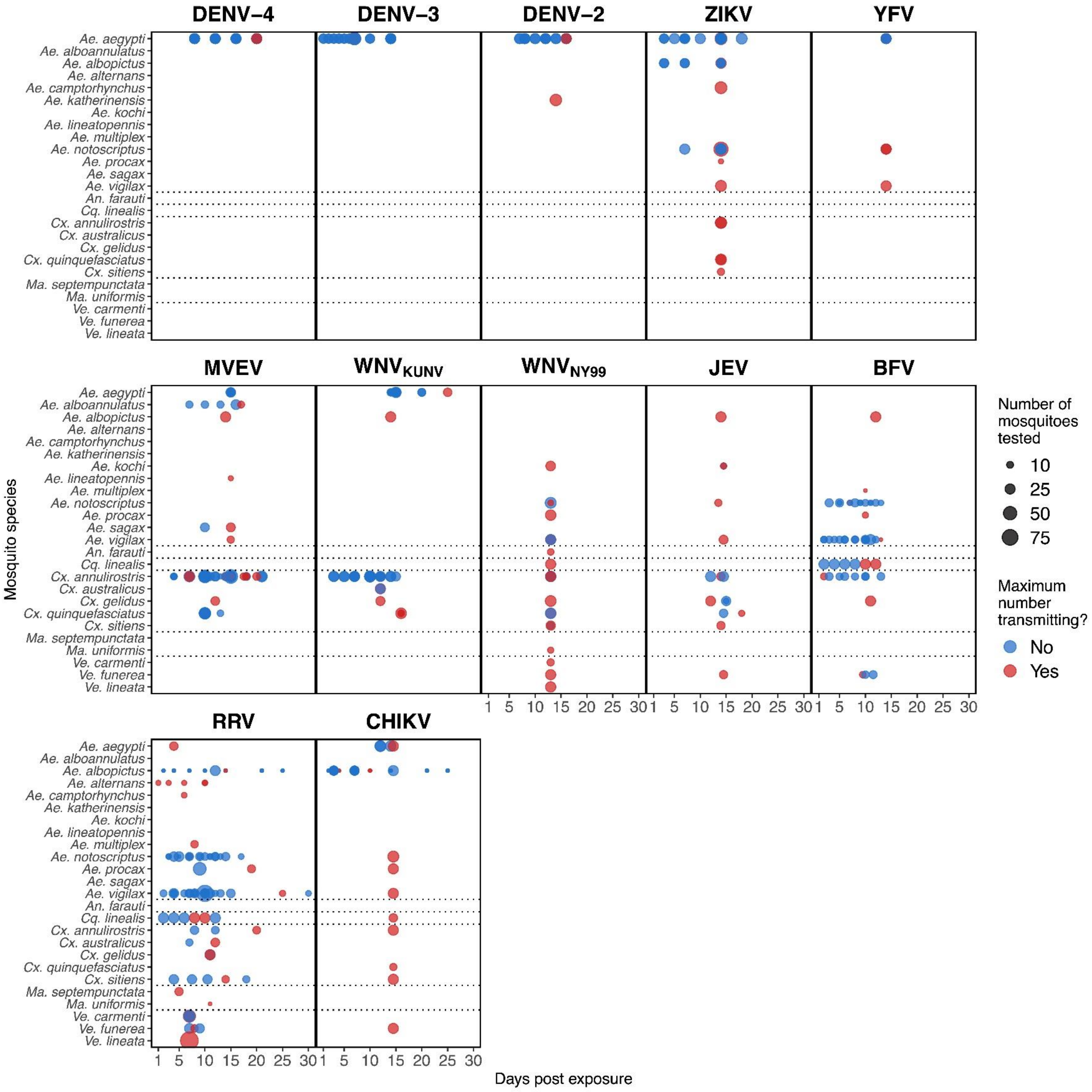


**S6 Fig**. **Days post viral exposure on which mosquitoes were tested for their ability to transmit viruses.** Blue points show experimental days that did not lead to the highest detected proportion of transmitting mosquitoes; red points show the day[s] that resulted in the highest observed transmitting proportion.

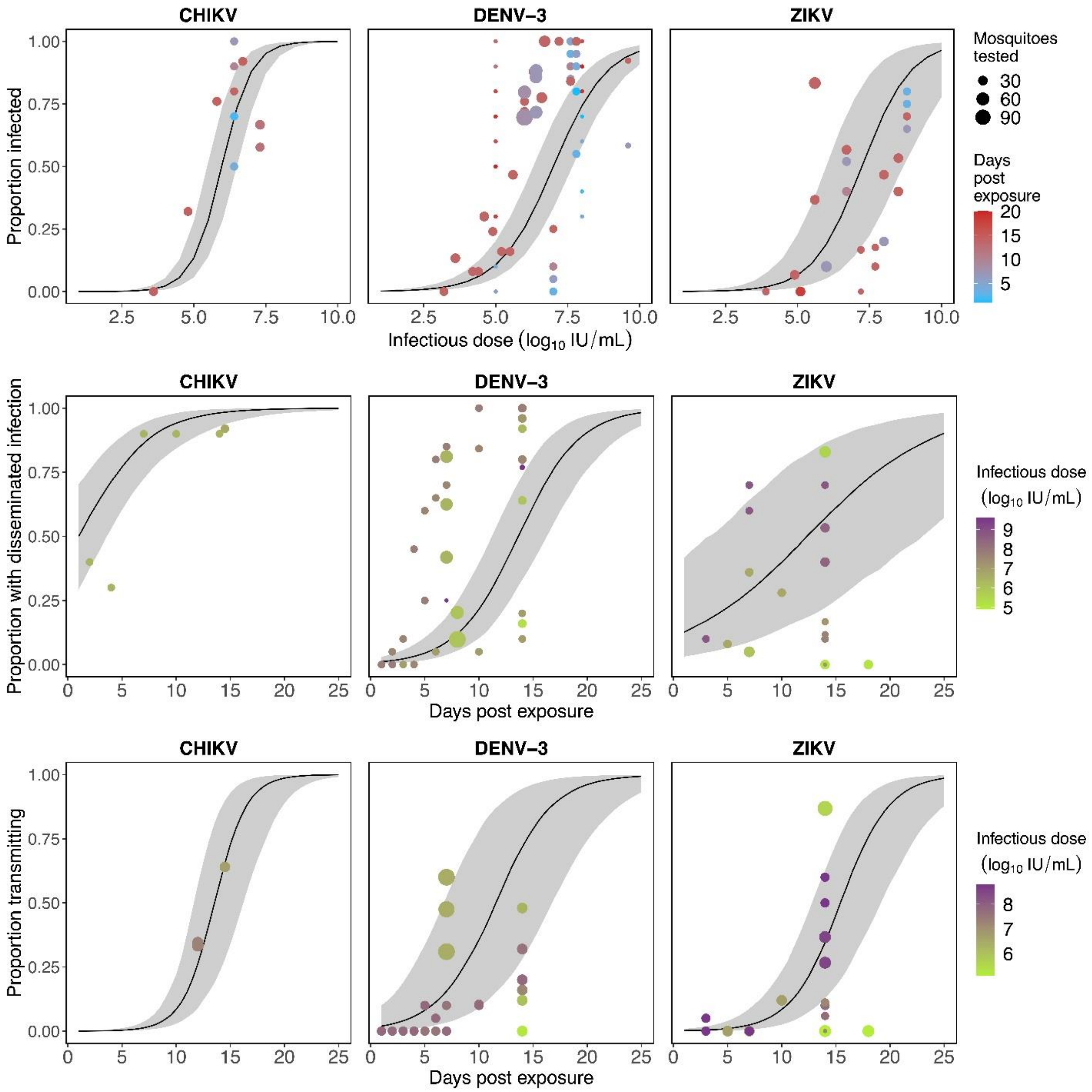


**S7 Fig. Continuous functions for mosquito infection over dose (top), and for dissemination and transmission over time (middle and bottom, respectively).** The results pictured here are from a generalized linear mixed effects model (GLMM) using *Aedes aegpyti* for all viruses they have been tested with in the lab; however, here we only show the three viruses for which more than one data point exist for infection, dissemination, and transmission. For infection (top), medians (solid lines) and 95% confidence intervals (grey bands) are estimated using twelve days post inoculation; for dissemination (middle) and transmission (bottom) estimates are drawn for a dose of 7 $\log_{10}$ IU/mL.
